# Effect of fortified balanced energy-protein supplementation during pregnancy and lactation on infant neurodevelopment: a community-based randomized controlled trial in rural Nepal

**DOI:** 10.64898/2026.07.02.26357185

**Authors:** Tsering P. Lama, Rita Shrestha, Parul Christian, James M. Tielsch, Joanne Katz, Subarna K. Khatry, Steven C LeClerq, Daniel J. Erchick

## Abstract

**Introduction:** Infant neurodevelopment is associated with maternal nutritional status, yet few studies have evaluated the effects of balanced energy–protein (BEP) supplementation during pregnancy and/or lactation.

**Methods:** The Maternal Infant Nutrition Trial (NCT03668977) was a 2×2 factorial, household randomized, unblinded, efficacy trial conducted in Sarlahi District, Nepal. Pregnant women were randomized to daily fortified BEP supplementation or no supplementation, and those with a live birth were re-randomized after delivery to supplementation or not until 6-months postpartum. The first 100 infants in each of the four intervention groups were enrolled in a neurodevelopment substudy. At 6-months, these infants underwent a cognitive, motor, language, and socioemotional assessment using the Bayley Scales of Infant and Toddler Development, fourth edition.

**Results:** Baseline characteristics were similar across the four trial groups. A significant interaction (p< 0.05) between pregnancy and lactation supplementation groups was observed in the language domain only. Infants born to mothers who received BEP supplementation during pregnancy did not differ in standard scores for cognitive (mean difference (MD): 2.0, 95% confidence interval (CI): -0.29, 4.29), motor (MD: 0.98, 95% CI: -1.41, 3.36), or socioemotional (MD: -2.35, 95% CI: -5.27, 0.57) domains compared to the pregnancy control group. Similarly, for these three domains, there were no significant differences among infants whose mothers received BEP in lactation period vs. not. Infants born to mothers supplemented in both pregnancy and lactation had significantly higher mean language standard scores of 2.12 (95% CI: 0.16, 4.08; p-interaction = 0.015), driven by the expressive communication subtest, compared to those with no BEP supplementation in either period.

**Conclusion:** BEP supplementation during pregnancy and/or lactation was not associated with improvements in cognitive, motor, or socioemotional domains at 6 months; however, infants of women supplemented in pregnancy and lactation had significant improvements in the language domain. Evaluation at later ages is warranted.

## Introduction

An estimated 250 million children under the age of five in low- and middle-income countries (LMICs) fail to reach their full cognitive potential due to poverty, poor health and nutrition, and limited access to and quality of care, increasing their risk for diminished educational outcomes and reduced future economic achievement (1). Maternal nutrition is a potentially modifiable factor important for fetal neurodevelopment, and nutritional status during pregnancy is linked to fetal development at all stages, including brain development, which in turn affects behavior and cognitive function (2). Suboptimal prenatal nutrition and maternal stress may disrupt key developmental processes, such as synaptic plasticity, neurogenesis, and dendritic arborization (3). The role of certain nutrients in the ontogenetic development of the central nervous system is well established, especially iron, n-3 fatty acids, and folate (4,5).

The World Health Organization (WHO) has recommended the use of balanced energy-protein (BEP) dietary supplementation during pregnancy in undernourished populations in their 2016 antenatal care guidelines (6). This recommendation is based on findings from systematic reviews of BEP supplementation during pregnancy that demonstrate moderate levels of evidence that such dietary supplementation can decrease the risk of stillbirth and SGA births and increase birth weight (7–9). However, due to the variation in type of BEP supplements, the Gates Foundation convened an expert group in 2016 to recommend the optimal nutritional composition of a BEP supplement, including micronutrients and calcium, for use in pregnant and lactating women in low income and food insecure contexts (10). Subsequently, several independent randomized controlled trials (RCTs) were conducted to evaluate the efficacy of the newly aligned fortified BEP supplement formulation when provided during pregnancy and/or lactation. These trials used harmonized designs and outcomes to enable pooled analyses of the effects of fortified BEP supplementation on birth outcomes, infant growth, and broader maternal and child health outcomes (11,12). The trials examining BEP provided to lactating women report modest improvements in infant growth (e.g., at 6 months) but currently show no clear evidence for direct, measurable gains in infant neurodevelopment by standard early measures (13–15). Neurodevelopment in children is also strongly influenced by early life experiences, including nutrition, illness and inflammation, and stress, particularly in infancy (16). The first six months of life are a period of rapid brain development, when many infants are solely reliant on human milk for their nutrition (17). Human milk contains several micronutrients, including vitamins B6, B12, choline, iodine, and selenium, among others, which are essential for neurodevelopment, including neurogenesis, differentiation, myelination, and neurotransmitter concentration (16,18). Importantly, the concentrations of many of these nutrients in milk are modifiable through maternal dietary intake (19). A meta-analysis of prenatal and postnatal nutrition intervention on mental development of children under two years in LMICs showed postnatal supplements consisting of macronutrients yielded a small effect on mental development but a null effect for prenatal intervention (20).

Collectively, findings suggest that adequate maternal nutrient intake, particularly involving both macro- and micronutrients may play a critical role in supporting early cognitive, language, and motor development. However, there are important gaps related to the efficacy of prenatal and postpartum macronutrient and micronutrient supplementation and neurodevelopment of children in LMICs. This study examined the effect of a fortified BEP supplement during pregnancy and/or for 6 months of lactation on infant neurodevelopment at 6 months of age.

## Methods

### Study design and participants

The Maternal and Infant nutrition Trial (MINT) was a 2×2 factorial, household randomized, unblinded, efficacy trial of daily fortified BEP supplementation vs. no supplementation among women during pregnancy and lactation (21). The trial was conducted from November 2021 to September 2024 in Sarlahi District, in the Terai region (low-lying plains) of Nepal. The primary outcomes were incidence of small-for-gestational age (SGA) birth and infant mean length for age z-score at 6 months of age. Infant neurodevelopment at age 6-months was a secondary outcome of the trial.

Sarlahi District, located in Madhesh Province, faces a disproportionate burden of intergenerational undernutrition, with stunting (29%) and underweight (27%) among children under five, exceeding national averages (25% and 19%, respectively), alongside the highest provincial proportions of women aged 20–49 with short stature (<145 cm; 13%) and low BMI (<18.5 kg/m²; 19%) in Nepal (22). The study area comprised 34 wards of Sarlahi District, selected from four municipalities and two rural municipalities, consisting of population of approximately 100,000 people.

A household census conducted in July 2019 (updated December 2022–January 2023) identified eligible married women aged 15–30 years, who were visited every five weeks by local female data collectors for pregnancy surveillance, with pregnant women reporting no allergy to nuts, milk, or soy enrolled in two waves (November 2021–July 2022 and January 2023–August 2023). A total of 1,944 pregnant women were enrolled in the trial, including 938 women in the first and 1,006 in the second enrollment period. Assessments for the current study were conducted in a subsample of infants born to women enrolled in the first enrollment period. All women and their infants who turned 6 months of age after November 21, 2022, were enrolled in the neurodevelopment substudy until the sample size was met.

### Ethics statement

The study received ethical approval from the Institutional Review Board (IRB) at Johns Hopkins Bloomberg School of Public Health, Baltimore, USA (IRB00009714), the Committee on Human Research IRB at The George Washington University, Washington, DC, USA (081739), and the Ethical Review Board of the Nepal Health Research Council, Kathmandu, Nepal (174/2018). The trial is registered at clinicaltrials.gov (NCT03668977). A data and safety monitoring board was established to review the study har and procedures prior to the start of the trial and monitor progress, safety, and findings during data collection. Written informed consent was obtained for participation in the trial from all women prior to enrollment and additional informed consent for participation in the substudy was obtained from the subset of women selected for the neurodevelopment substudy. Married women under 18 years of age were considered emancipated minors and required an additional written consent from both the woman and her guardian (who is an adult) when obtaining consent. An assent form was not required by the IRB.

### Randomization

After obtaining informed consent, eligible pregnant women were randomized at the household level to either the daily fortified BEP supplementation or not. Randomization was stratified by maternal height (<150 cm, ≥150 cm), based on the median of three measurements, using pre-generated permuted blocks (sizes 4, 6, 8, and 12) to ensure group balance. We pre-generated randomization sequences using the R software environment for statistical programming (https://www.r-project.org/) and the tablets that were used for data collection obtained the random allocation based on the pre-generated sequence that is stored in the central server. At the birth visit, postpartum randomization was conducted among households with at least one live birth, stratified by maternal height and pregnancy allocation. If a household already had an assigned allocation code from a prior participant, the same code was used for any additional participants enrolled in that household. The study personnel who enrolled and assigned participants to the intervention did not have access to the random allocation sequence. The study followed a 2×2 factorial design with four groups: (i) no BEP supplementation during pregnancy and lactation (control), (ii) BEP supplementation during pregnancy only, (iii) BEP supplementation during lactation only, and (iv) BEP supplementation during both pregnancy and lactation. Due to the nature of the intervention, the participants and study data collectors were not blinded to the intervention.

### Intervention

Women in the pregnancy and/or lactation supplement groups received fortified BEP supplements for daily consumption. The daily fortified BEP supplement was a lipid-based peanut paste in a single sachet serving of 72 g (∼400 kcal; 14.5-g protein) fortified with 15 multiple micronutrients provided at an recommended dietary allowance for pregnancy and 500 mg of calcium and designed to meet the expert consensus specification for nutritional requirements during pregnancy and lactation (10). This product is a variation of ‘Plumpy’Mum’ produced by Nutriset (France) for the purpose of research. This BEP supplement was selected for the trial after two rounds of formative research in the same study area that assessed acceptability and compliance of various forms of BEP supplements (23,24).

Both intervention and control groups received the following: 1) recommendation to enroll at the local health post for ANC and deliver at a health facility with a birthing facility; 2) counseling on nutrition, hygiene, breastfeeding and newborn care; 3) clean birthing kit and 4) provision of iron folic acid tablet (IFA) if not provided at ANC by the health facility. At the birth visit, the women were randomly allocated to postpartum BEP supplementation or not, and women in both postpartum groups received breastfeeding counseling.

### Trial procedures

Following enrollment, participants underwent a baseline visit and ultrasound examination between 10 and 24 weeks of gestation to assess gestational age using a portable machine (SonoSite NanoMaxx). Women in both pregnancy groups were visited weekly at home by data collectors from 14 weeks’ gestation until pregnancy outcome for monitoring of pregnancy morbidities, with women in the supplement group additionally receiving BEP supplements and counseling. A late pregnancy visit (32–34 weeks’ gestation) was conducted to collect data on maternal anthropometry, pregnancy morbidities, and delivery plans. Families were instructed to notify the local data collector at labor onset to enable the birth assessment within 72 hours of delivery. During the birth visit, data collectors conducted home visits to collect anthropometric measurements of the mother and infant, along with information on the birth and delivery process, early breastfeeding and feeding practices, and newborn care. Women in both lactation groups were visited weekly for monitoring maternal and infant health and providing the BEP supplements in the intervention group. Additionally, monthly visits were conducted up to 6 months postpartum to collect data on maternal and infant morbidity, feeding and care practices, household food security, and anthropometric measurements of both mother and infant. A final visit at 12 months postpartum was conducted to assess infant growth as a secondary outcome among a subsample prior to administrative end of the study. For those enrolled in the neurodevelopment substudy, one of two trained psychologists conducted the assessment at the nearest field site in the presence of the primary caregiver.

### Neurodevelopment assessments and outcomes

Neurodevelopmental outcomes at 6 months of age were secondary outcomes of this trial. The Bayley Scales of Infant and Toddler Development, fourth edition (Bayley-4), was used to measure infant functioning in four domains: cognition, motor (fine motor and gross motor), language (receptive and expressive language), and socio-emotional. The Bayley-4 cognitive domain assesses infants’ interest in novelty, attention to familiar and unfamiliar objects, and play behavior. The language domain includes receptive and expressive communication subtests: receptive tasks evaluate recognition of sounds, objects, and people in the environment, while expressive tasks capture the use of sounds, gestures, and nonverbal cues (e.g., smiling, vocalizing, laughing) within social contexts. The motor domain in infants covers fine motor skills (visual tracking, hand-to-mouth, object transfer, grasping) and gross motor skills (head control, rolling, sitting, and crawling). Finally, the socio-emotional domain tracks age-expected milestones in social interaction and responses to sensory input such as sound and touch (25). Bayley-4, a commonly used assessment of infant development, was selected for this study because the instrument has been adapted to the Nepali cultural context and allows for comparison with data from previous studies conducted in Nepal. The previous version, Bayley-III, was adapted for use in a study of Nepali children in Bhaktapur District, and had high inter-rater, moderate to high internal consistency, and distributions of cognitive and motor tests comparable to a standard population, supporting its feasibility for assessing infant neurodevelopment in the Nepali cultural context (26). The questionnaire-based social-emotional subtests were translated using the previously tested and adapted version used in the Malnutrition and Enteric Diseases (Mal-ED) study in Bhaktapur district, Nepal (27,28). The Bayley-4 was pilot tested among 6-month-old children in Sarlahi not enrolled in the trial. Study psychologists and a senior psychologist, serving as the gold standard, independently scored the assessments, demonstrating high inter-rater reliability with minimal inter-observer differences.

In this trial, Bayley-4 assessments were conducted by two trained psychologists supervised by the senior consultant psychologist. The senior consultant psychologist oversaw quality assurance/quality control, training, supervision and monitoring. Data collection for the neurodevelopment substudy started on November 21, 2022, and ended on April 25, 2023. At the 6-month postpartum home visit, mothers provided consent for their infants to undergo neurodevelopmental assessment, and we enrolled 400 infants (100 from each intervention arm). Assessments were conducted at field offices by the two psychologists working simultaneously at separate sites, with dates scheduled based on participant availability. Infants were transported with their caregiver by project vehicle and assessed only if healthy and well-fed; otherwise, testing was rescheduled. In line with Bayley-4 standards, cognitive, language (receptive and expressive), and fine motor skills were assessed with the infant seated on the caregiver’s lap, while gross motor skills were evaluated on a floor mat.

To ensure inter-observer reliability, 10% of the Bayley-4 assessments were double-scored by a consultant psychologist using video recordings of the sessions. Each assessment lasted approximately one hour. Data were first recorded on paper forms and then entered into Pearson’s Q-global web-based system for scoring and reporting (29). Raw scores for each domain at 6 months were converted into scaled scores with a mean of 10 and SD of 3, and a range of 1-19 for each domain and subtest. The scaled scores were converted to a standard score equivalent scores, scaled from 45 to 155, with a mean of 100 and a standard deviation of 15 where scores of 85 and 115 are 1 standard deviation (SD) below and above the mean, respectively (25). A standard score of 76-85 is classified as low average, 86-109 score is average, 110-119 standard score is high average, 120-129 is very high, and above 129 classified as extremely high. In addition, the two psychologists also conducted the WHO six gross motor milestones module, which assessed the following: sitting without support, hands-and-knees crawling, standing with assistance, walking with assistance, standing alone and walking alone (30).

### Sample size

The sample size for this sub-study was calculated to detect a difference of 0.58 in scaled cognitive scores between the pregnancy supplementation vs. not groups, assuming no interaction between the pregnancy and postpartum interventions. A sample size of 200 infants per pregnancy intervention arm (400 total), assuming a mean scaled cognitive score of 9.55 and a standard deviation of 2.05, provided adequate power to detect this difference. Given the trial’s 2×2 factorial design, the target sample was further stratified by postpartum intervention, resulting in approximately 100 infants per combined pregnancy–postpartum intervention group.

### Statistical analysis

For the domains with only one subtest (i.e., cognitive and socio-emotional), the scaled score to standard score equivalent is a linear conversion from one scale (mean=10, SD=3) to another (mean=100, SD=15) (25). For the domains with two subtests (i.e. language and motor), the distribution of the sum of scaled scores was converted to a scale with a mean of 100 and SD of 15. The raw, scaled, and standard scores were generated based on the age group as part of the individual assessment report by Bayley.

The inter-rater reliability between the two psychologists and the senior psychologists assessed for quality control was expressed using the intraclass correlation coefficient (ICC). Correlations between the raw and scaled scores of the different subtests were assessed.

For this analysis, the standard score for each scale was used as the outcome variable. The baseline characteristics of each of the four randomized groups were compared for balance across. The interaction between BEP in pregnancy and BEP in lactation on neurodevelopmental outcomes was assessed for each Bayley-4 domains. Multivariable linear regression model was used to estimate the mean difference and 95% confidence intervals for each of the four neurodevelopmental outcome domains, with the group that did not get the intervention serving as the reference. For domains with a significant interaction (p < 0.05), a regression model including both the main effects and the interaction term was fitted. For domains with no evidence of interaction, a main-effects model was estimated to assess the marginal effects of the BEP supplementation in pregnancy and BEP supplementation in lactation separately.

A bivariate analysis was conducted to assess differences in the achievement of the various WHO gross motor skill indicators by the four groups. Analyses were conducted in Stata version 15 (31).

## Results

The neurodevelopment substudy was conducted among the infants born to women enrolled in the first cohort of the trial. At that time, 8,370 women were eligible for pregnancy surveillance, 999 pregnancies were detected, and 938 women were enrolled in the first cohort (Figure 1). Study participation flowchart for the neurodevelopment sub-study

The baseline characteristics were evenly distributed between the intervention groups (Table 1a and 1b). The mean age at the time enrollment was 24.4 (SD 3.4). Nearly half (46%) of women had no schooling, 6% of the women were nulliparous, and 31% of the women were underweight (<18.5kg/m^2^) in this group. Most characteristics of participants whose children had neurodevelopmental assessments were like those who did not (S1 Table).

**Figure 1.**
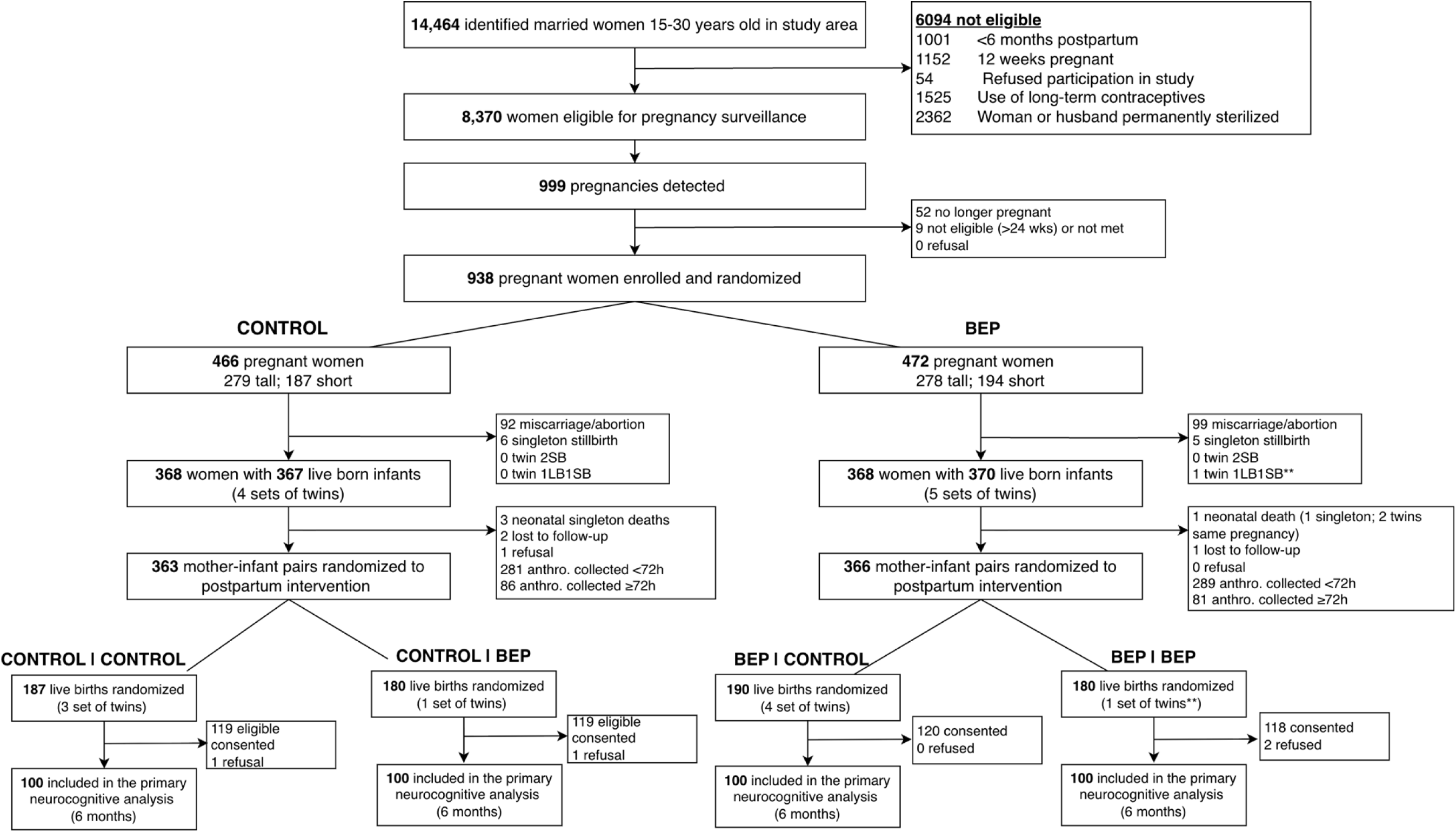
Study participation flowchart for the neurodevelopment sub study.

**Table 1a.** Baseline characteristics of participants in the neurodevelopment substudy (n=400)

|  | **Control in pregnancy and lactation** | **BEP in pregnancy, control in lactation** | **Control in pregnancy, BEP in lactation** | **BEP in pregnancy and lactation** |
| --- | --- | --- | --- | --- |
| **Characteristics, %** | **n=100** | **n=100** | **n=100** | **n=100** |
| **Age at baseline (years)** | **n (%)** | **n (%)** | **n (%)** | **n (%)** |
| <20 | 3 (3.0) | 6 (6.0) | 3 (3.0) | 3 (3.0) |
| 20-24 | 51 (51.0) | 50 (50.0) | 52 (52.0) | 52 (52.0) |
| 25-29 | 38 (38.0) | 35 (35.0) | 28 (28.0) | 38 (38.0) |
| 30 or more | 8 (8.0) | 9 (9.0) | 17 (17.0) | 7 (7.0) |
| **Mean age of first marriage (years)** | 17.6 ± 2.2 | 17.6 ± 1.9 | 17.4 ± 2.0 | 17.3 ± 2.2 |
| **Parity** |  |  |  |  |
| Nulliparous | 9 (9.0) | 4 (4.0) | 5 (5.0) | 6 (6.0) |
| 1 | 40 (40.0) | 46 (46.0) | 39 (39.0) | 34 (34.0) |
| 2 | 31 (31.0) | 27 (27.0) | 31 (31.0) | 33 (33.0) |
| ≥3 | 20 (20.0) | 23 (23.0) | 25 (25.0) | 27 (27.0) |
| **Years of education** |  |  |  |  |
| No Schooling | 43 (43.0) | 36 (36.0) | 50 (50.0) | 54 (54.0) |
| 1-5 years (Primary) | 15 (15.0) | 19 (19.0) | 23 (23.0) | 13 (13.0) |
| 6-10 years (Junior Secondary) | 23 (23.0) | 24 (24.0) | 13 (13.0) | 17 (17.0) |
| 11-16 years (Higher secondary) | 19 (19.0) | 21 (21.0) | 14 (14.0) | 16 (16.0) |
| **Ethnicity** |  |  |  |  |
| Pahadi | 2 (2.0) | 3 (3.0) | 0 (0.0) | 0 (0.0) |
| Madeshi | 98 (98.0) | 97 (97.0) | 100 (100.0) | 100 (100.0) |
| **Religion** |  |  |  |  |
| Hindu | 77 (77.0) | 75 (75.0) | 78 (78.0) | 71 (71.0) |
| Muslim | 22 (22.0) | 24 (24.0) | 22 (22.0) | 29 (29.0) |
| Buddhist | 1 (1.0) | 1 (1.0) | 0 (0.0) | 0 (0.0) |
| **Baseline anthropometric measurements** |  |  |  |  |
| **BMI category (kg/m2)** |  |  |  |  |
| Underweight (BMI <18.5 ) | 32 (32.0) | 26 (26.0) | 37 (37.0) | 29 (29.0) |
| Normal weight (BMI 18.5- 24.9) | 63 (63.0) | 66 (66.0) | 59 (59.0) | 64 (64.0) |
| Overweight (25.0- 29.9) | 4 (4.0) | 8 (8.0) | 4 (4.0) | 5 (5.0) |
| Obese ($\geq 30.0$) | 1 (1.0) | 0 (0.0) | 0 (0.0) | 2 (2.0) |
| Mean BMI ± SD, kg/m<sup>2</sup> | 20.3 ± 2.9 | 20.4 ± 2.9 | 20.0 ± 2.6 | 20.4 ± 3.2 |
| Mean weight ± SD, kg | 45.9 ± 7.4 | 47.4 ± 7.8 | 45.3 ± 6.5 | 46.4 ± 8.0 |
| Mean height ± SD, cm | 150.4 ± 5.1 | 152.0 ± 5.1 | 150.3 ± 5.0 | 150.8 ± 5.2 |
| Height <150 cm | 47 (47.0) | 35 (35.0) | 44 (44.0) | 41 (41.0) |
| Height $\geq 150$ cm | 53 (53.0) | 65 (65.0) | 56 (56.0) | 59 (59.0) |
| Mean MUAC ± SD, cm | 24.4 ± 2.5 | 24.5 ± 2.8 | 24.2 ± 2.5 | 24.4 ± 2.7 |
| MUAC <23.0 cm | 33 (33.0) | 29 (29.0) | 34 (34.0) | 29 (29.0) |
| MUAC $\geq 23.0$ | 67 (67.0) | 71 (71.0) | 66 (66.0) | 71 (71.0) |
| **Wealth Quintile** |  |  |  |  |
| Lowest wealth quintile (poorest) | 19 (19.0) | 12 (12.0) | 18 (18.0) | 24 (24.0) |
| Second wealth quintile | 21 (21.0) | 22 (22.0) | 17 (17.0) | 24 (24.0) |
| Middle wealth quintile | 27 (27.0) | 18 (18.0) | 22 (22.0) | 16 (16.0) |
| Fourth wealth quintile | 19 (19.0) | 20 (20.0) | 20 (20.0) | 18 (18.0) |
| Highest wealth quintile (richest) | 14 (14.0) | 28 (28.0) | 23 (23.0) | 18 (18.0) |
| **Household food insecurity** |  |  |  |  |
| Food secure | 76 (76) | 78 (78) | 74 (74) | 66 (66) |
| Mild | 12 (12) | 7 (7) | 14 (14) | 9 (9) |
| Moderate | 7 (7) | 12 (12) | 12 (12) | 20 (20) |
| Severe | 5 (5) | 3 (3) | 0 (0) | 5 (5) |

**Table 1b.** Characteristics of the infants, delivery practices, and breastfeeding practices in the neurodevelopment substudy.

| **Delivery and infant characteristics** | **Control in pregnancy and lactation**<br>**n=100**<br>**n (%)** | **BEP in pregnancy, control in lactation**<br>**n=100**<br>**n (%)** | **Control in pregnancy, BEP in lactation**<br>**n=100**<br>**n (%)** | **BEP in pregnancy and lactation**<br>**n=100**<br>**n (%)** |
| --- | --- | --- | --- | --- |
| **Child delivery variables** |  |  |  |  |
| **Delivery type** |  |  |  |  |
| Normal | 86 (86.0) | 93 (93.0) | 93 (93.0) | 86 (86.0) |
| C-section | 14 (14.0) | 7 (7.0) | 7 (7.0) | 14 (14.0) |
| **Delivery location** |  |  |  |  |
| Home | 15 (15.0) | 24 (24.0) | 18 (18.0) | 15 (15.0) |
| Maiti (parental) home | 6 (6.0) | 1 (1.0) | 2 (2.0) | 6 (6.0) |
| Health post/clinic | 44 (44.0) | 45 (45.0) | 56 (56.0) | 50 (50.0) |
| Hospital | 30 (30.0) | 28 (28.0) | 22 (22.0) | 27 (27.0) |
| On the way to facility | 5 (5.0) | 2 (2.0) | 2 (2.0) | 2 (2.0) |
| **Infant variables** |  |  |  |  |
| **Sex (%)** |  |  |  |  |
| Male | 53 (53.0) | 58 (58.0) | 50 (50.0) | 61 (61.0) |
| Female | 47 (47.0) | 42 (42.0) | 50 (50.0) | 39 (39.0) |
| **Breast feeding history** |  |  |  |  |
| Breastfeeding within 1 hour of birth | 50 (50.0) | 57 (57.0) | 61 (61.0) | 55 (55.0) |
| **Breastfeeding at 6 months visit** | **n=93** | **n=90** | **n=83** | **n=92** |
| Exclusive breastfeeding for 6 months | 15 (16.1) | 23 (25.6) | 22 (26.5) | 21 (22.8) |

### Effect of the Interventions on Neurodevelopmental Outcomes

There was no evidence of interaction between BEP supplementation in pregnancy and lactation for cognitive, motor, and socioemotional domains. However, a significant interaction between supplementation periods was identified for the language domain (p-value=0.015) (S3 Table). Tests of interaction conducted for the two subtests of the language domain (i.e., receptive and expressive scaled scores) indicated that there was a significant interaction only for the expressive subtest only (p-value= 0.008) (data not shown).

### Marginal effects of BEP supplementation in pregnancy and lactation

The individual effects of BEP supplementation during pregnancy or lactation on the cognitive, motor, and socioemotional domain are shown in Table 2a. Compared with the no BEP in pregnancy group, infants born to mothers who received BEP supplements during pregnancy did not differ in standard scores for the cognitive (mean difference [MD]: 2.0, 95% CI: -0.29, 4.29), motor (MD: 0.98, 95% CI: -1.41, 3.36), or socioemotional (MD: -2.35, 95% CI: -5.27, 0.57). Similarly, for these three domains, there were no significant differences among infants born to mothers who received BEP in lactation vs. who did not receive BEP in lactation period (Table 2a).

**Table 2a.** Neurodevelopmental outcome (cognitive, motor and socio-emotional) mean standard scores and difference by intervention group (marginal effects)

| *Outcomes* | *Mean (SD) standard score* | *Mean (SD) standard score* | *MD<sup>1</sup> (95%CI)* | *Mean (SD) standard score* | *Mean (SD) standard score* | *MD<sup>1</sup> (95%CI)* |
| --- | --- | --- | --- | --- | --- | --- |
|  | No BEP in pregnancy (a) | BEP in pregnancy (b) | BEP in pregnancy vs. No BEP in pregnancy (b)-(a) | No BEP in lactation (c) | BEP in lactation (d) | BEP in lactation vs. No BEP in lactation (d)-(c) |
| **Cognitive** | 97.5 (11.7) | 99.5 (11.6) | 2.0 (-0.29, 4.29) | 98.5 (10.9) | 98.5 (12.5) | -0.05 (-2.35, 2.25) |
| **Motor** | 104.3 (11.9) | 105.2 (12.3) | 0.98 (-1.41, 3.36) | 105.2 (11.8) | 104.3 (12.5) | -0.91 (-3.30, 1.47) |
| **Socio-emotional** | 130.1 (14.6) | 127.8 (15.1) | -2.35 (-5.27, 0.57) | 128.3 (14.7) | 129.6 (15.1) | 1.3 (-1.63, 4.22) |

SD, standard deviation; MD, mean difference; 95% CI, 95% confidence interval
<sup>1</sup> Multivariable linear regression analysis without the interaction effect

### Combined effect of BEP supplementation in pregnancy and lactation

Infants born to mothers in the group supplemented in both pregnancy and lactation periods had significantly higher mean language standard scores of 2.12 standard scores (95% CI: 0.16, 4.08), driven by the expressive communication subtest, compared to women in the group with no supplementation in either period (Table 2b).

**Table 2b.** Neurodevelopmental language outcome mean scores and difference by intervention group (interaction effect)

| *Outcome* | *Mean standard score (SD) by group* |  |  |  | *Mean Difference<sup>1</sup> (95% CI)* |  |  |
| --- | --- | --- | --- | --- | --- | --- | --- |
|  | Control in pregnancy and lactation (a) | BEP in pregnancy, control in lactation (b) | Control in pregnancy, BEP in lactation (c) | BEP in pregnancy and lactation (d) | BEP in pregnancy only vs control group (b)-(a) | BEP in lactation only vs control group (c)-(a) | BEP in pregnancy and lactation vs control group (d)-(a) |
| **Language** | 98.48 (7.13) | 97.99 (7.22) | 97.64 (6.99) | 100.60 (6.87) | -0.49 (-2.45, 1.47) | -0.84 (-2.80, 1.12) | **2.12 (0.16, 4.08)** |

SD, standard deviation; 95% CI, 95% confidence interval
<sup>1</sup> Multivariable linear regression analysis with the interaction effect

### Gross motor milestones

The WHO Gross motor milestones assessment at 6 months of age showed that only about 21% of infants were able to sit without support and this did not vary significantly by the four groups (S4 Table). Less than 3% of the infants were able to crawl on their hands and knees. Most children were awake and alert during the assessment.

## Discussion

In this community-based randomized controlled trial in rural Nepal, daily fortified BEP supplementation in pregnancy or lactation did not improve cognitive, motor, or socioemotional functioning among infants 6 months of age. However, combined BEP supplementation in pregnancy and lactation improved infant language scores by an average of 2.12 standard scores (95% CI: 0.16, 4.08), driven by the expressive communication subtest.

The MINT study may not be directly comparable to other studies, as few have evaluated similar fortified BEP interventions or assessed neurodevelopment at 6 months of age. In an integrated RCT of health, nutrition (micronutrient and nutrition protein rich food supplement), psychosocial, and water, sanitation, and hygiene interventions delivered from preconception, pregnancy through early childhood in India, children in the intervention arm demonstrated significantly higher cognitive, language, motor and socioemotional scores at 24 months compared with controls (32). Notably, combined interventions across all three periods (preconception, pregnancy and early childhood) yielded the greatest gains, while preconception interventions alone had limited impact, improving only cognitive scores (32). Unlike this trial in India, our trial was focused on nutrition supplementation only and did not have a preconception intervention arm nor provided complementary feeding to infants after 6 months of age.

Other randomized controlled trials have further explored the effects of prenatal nutrient supplementation on child development. A recent trial conducted in rural Niger comparing multiple micronutrient supplements (MMS) and medium-quantity lipid-based nutrient supplements (LNS) fortified with MMS compared to routine IFA supplementation found that prenatal LNS improved cognitive development trajectories at 18 and 21 months, though effects were not sustained at 24 months (33). The same study reported earlier attainment of sitting and walking milestones among children of mothers who received LNS compared with those who received IFA. An RCT in Bangladesh that compared the effects on infant development of early (8 –10 weeks’ gestation) or later (about 17 weeks’ gestation) supplementation with food (600kcal of energy) and multiple micronutrients, 30 mg Fe 400 g folate, or 60 mg Fe 400 g folate found that the prenatal food and multiple micronutrient supplementation had no significant effect on infant development at 7 months (34). Small benefits in problem-solving, motor scores, and activity levels were only observed among infants of undernourished mothers (BMI < 18.5 kg/m^2^), though the authors noted these effects were of doubtful functional importance (34). A previous cohort follow-up of 676 children aged 7 to 9 years, born to women in 4 of 5 groups of a RCT of prenatal micronutrient supplementation (IFA, IFA with zinc, 11 micronutrients vs. control group Vitamin A only) in a population similar to our trial in rural Nepal, showed that only the prenatal IFA offspring had better intellectual and motor function than the control group (35). The China National Birth Cohort showed that maternal adherence to a high-protein and micronutrient-rich dietary pattern during pregnancy was associated with better neurodevelopmental outcomes such as higher gross motor and problem solving skills at 36 months of age (36). An RCT in Vietnam where pregnant women in the intervention group received maternal milk supplementation consisting of two daily servings of supplementation (which provided 252 kcal, 16.8 g protein, 1.4 g fat, 39.2 g carbohydrate, and a variety of micronutrients) with a breastfeeding support program from late pregnancy to 12 weeks lactation found that children born to mothers in the intervention group had significantly higher cognitive and motor functions at 30 months of age (37). A systematic review of 18 RCTs with heterogeneous measures found no conclusive overall effect of prenatal micronutrient supplements on children’s mental development; there were signals of benefit for multi-micronutrients (and n-3 fatty acids) in some domains, but evidence was inconsistent and trials/methods varied widely (38). Another meta-analysis found that the timing of prenatal supplementation when implemented in the first trimester showed improved cognitive development in children (39).

Evidence on the effect of nutrition supplementation given in lactating period to mothers on child neurodevelopment were lacking with most postnatal supplementation RCTs provided to infants as early as two months after birth (20). The effects of nutrition intervention on child development may be mediated through improved birth outcomes, postnatal growth, or breastfeeding practices. In our trial, BEP supplementation did not improve birth or growth outcomes (*publication under review*), and the prevalence of exclusive breastfeeding was suboptimal (38-44 %), which may partly explain the lack of observed effects on early child development.

A meta-analysis of prenatal micronutrient supplementation showed evidence that interventions with ≥5 years of follow-up (i.e., the time between intervention completion and outcome assessment) was significantly associated with cognitive benefits (39). Most studies of nutritional interventions in pregnancy conducted assessments at later ages, typically between 18 and 24 months (32,33,36,37,37,40). Neurodevelopment was assessed only at 6 months of age in our study, providing a limited snapshot of development in early infancy; the absence of follow-up assessments at later ages precluded examination of the developmental trajectory of neurocognitive functions. Neurodevelopment assessments show low reliability in early childhood (41–43); thus, assessments later in childhood (e.g., during the first years of school) could give more reliable and detailed information on the effect of the intervention. A longer follow-up of the children beyond 6 months to assess cognitive development trajectories at 18, 24 months and later would be beneficial to understand the longer-term effects of BEP supplement. Such longitudinal assessments would also clarify whether early differences observed in the language domain predict subsequent variations in other cognitive domains.

An RCT assessing the effect of maternal vitamin B12 supplementation on child cognitive outcomes in South Indian children at 30 months of age also found significant association with the language domain driven by the expressive subtest (44). A cluster randomized trial of multiple micronutrient powder (MNP) in Indian preschool children showed that MNP fortification positively impacted expressive language only in low-quality preschools (45). It is unclear why expressive language and not receptive language has been impacted. Receptive and expressive language require different abilities and therefore can develop independently (25). Receptively, newborns turn their eyes and head in the general direction of sounds, with sound localization abilities improving over the first six months (46). During the first year, infants become attuned to the overall patterns and rhythms of speech in their native languages (47). Expressive communication begins with cooing around two months of age, followed by babbling composed of single consonant–vowel sounds at 4–5 months (48). By 7–8 months, infants begin to combine these sounds into longer sequences, and by around 10 months, they produce varied syllabic combinations. The observed effect limited to the expressive language subdomain among children whose mothers received BEP supplementation during pregnancy and lactation is not fully understood and may reflect an early stage in the developmental trajectory, making it premature to infer its long-term significance.

There are plausible explanations for why we did not observe significant effects of BEP supplementation on neurodevelopment, except for in the language domain. Two complementary mechanisms may explain the neurodevelopmental effects especially on the language domain of early nutrition supplementation: (1) direct enhancement of brain development through increased nutrient supply, and (2) indirect effects via interactions with the child’s environment and behavior (49).

First, improved maternal nutritional adequacy from both macro- and micronutrients in our BEP supplement among those who received it during both pregnancy and 6 months lactation may have supported brain development during gestation and infancy, which is a critical period of neurodevelopment. Evidence shows that nutrient deficiencies during pregnancy and lactation impair neuronal growth, myelination, and brain formation, leading to deficits in motor, memory, learning, and cognitive functions (49,50). Key nutrients essential for infant neurological development, such as vitamin A, B, folate, iodine, selenium and fatty acids, are higher in the breast milk of mothers with better maternal nutrition providing greater support for postnatal growth and development (51,52). At birth, the infant brain weighs about 350 g (∼10% of body weight), nearly doubling to ∼660 g by 6 months. By 1 year, it reaches ∼925 g, around 70% of the adult brain weight (1300–1400 g, ∼2% of body weight) (53). Optimal nutrition in the first year of life is essential for brain growth and function, as nutrients play key roles in neurogenesis, myelination, neurotransmitter synthesis, and synaptic plasticity (54). However, brain development is a complex process with different parts and functions of the brain developing at different times (55). Functionally, sensory pathways such as those for vision and hearing develop first during the infant’s first year of life, followed by the emergence of early language abilities, and subsequently by higher-order cognitive functions that continue to mature into adulthood (56). In the domain of language, neural indicators of learning appear remarkably early in development, and these early neural responses are predictive of later language performance in children (56). Thus, the age of development assessments may have contributed to detecting language domain difference at the early 6-month assessment and not the other domains.

Second, differences in how caregivers talk to and involve infants affect the trajectory of language development. Environments characterized by insufficient sensory and social stimulation disrupt neurodevelopmental processes analogous to those compromised by early nutrient deficiency, including diminished dendritic arborization and synaptic density (49). Empirical evidence indicates that nutritional inadequacy and insufficient environmental stimulation exert distinct yet interrelated influences on neurodevelopment through additive, interactive, and mediating pathways (49). Both nutrition and early learning/responsive caregiving interventions have been effective in promoting early childhood development (57), with meta-analyses showing larger effects for caregiving than for nutrition (58,59). This was also evident in the cluster randomized study of MNP in preschools in India, where MNP fortification had a positive effect on expressive language and to a lesser extent, inhibitory control and social–emotional development in low-quality preschools and null effect in high-quality preschools (45). The home environment, which may act as an unmeasured confounder influencing the association between the intervention and child neurodevelopmental outcomes, was not assessed in our study. Inclusion of data on the quality of the home environment could have provided additional insight into the observed differences in expressive language at 6 months of age.

The study had several limitations. The study was not powered to assess the combined effect of BEP supplementation in pregnancy and lactation on neurodevelopmental outcomes, nor to evaluate potential effect modification by maternal undernutrition and child growth status.

Compared to many of the child neurodevelopmental studies, our assessment was conducted at an early age, which may limit the ability to detect meaningful differences and does not capture developmental trajectories during the first two years of life (60). Bayley-4 scores from a single assessment time point should not be used alone to diagnose cognitive delay; instead, it is recommended that the outcome measures should be collected at least at 2–3 points during that time of rapid change and maturation to optimize sensitivity (61). Lastly, while we relied solely on the Bayley-4, the use of multiple methods is recommended to more comprehensively evaluate neurodevelopmental outcomes (61).

The study also had important strengths. A major strength of the trial was the 2×2 factorial design, which allowed us to examine the independent effects of BEP supplementation during pregnancy alone, lactation alone, and in combination. Conducting the study in rural Nepal provided valuable evidence from among a population of women at high risk of undernutrition and micronutrient deficiencies (62). Additional strength is the high-quality assessment of outcomes including the high inter-rater agreement on the neurodevelopment outcomes. Our substudy population was based on a population-based sample of infants and generalizable to that population. Finally, previous versions of the Bayley-4 have been successfully applied in Nepal and shown to be reliable and acceptable in similar settings (26).

Direct evidence on the effects of fortified BEP supplementation during pregnancy and lactation on early childhood neurodevelopment remains limited, making it difficult to draw definitive conclusions. A significant finding was observed for the combined effect of BEP supplementation during pregnancy and lactation on the language domain, suggesting that more research is needed on the combined effect of BEP supplementation on infant neurodevelopment development. Studies are needed to evaluate the effect of nutritional interventions on neurodevelopment outcomes at multiple timepoints beyond 6 months.

## Supporting information

**S1 Table: Baseline characteristics of the first cohort of women enrolled by the intervention group (n=729)**

**S2 Table: Inter-rater reliability between the two psychologists and the senior psychologists on the Bayley IV subtests for quality control (n=41)**

**S3 Table: Effect of BEP supplementation on Bayley-4 domain outcomes at 6 months of age with test of interaction**

**S4 Table: Distribution of gross motor milestones at 6 months by the four groups Acknowledgements**

We express our sincere thanks to the women and their families who participated in the MINT Study and especially to the families, mother and infants who participated in the neurodevelopment substudy in Sarlahi District, Nepal. Thank you to Nutriset, France, for producing and donating a portion of the fortified BEP supplements. Thank you to the two-psychologist Ms. Kriti Joshi and Ms. Sunee Shrestha who conducted the neurodevelopment assessments for the study. Thank you also the members of our data and safety monitoring board, including Dr. Sudha Basnet (Chairperson), Department of Pediatrics, Institute of Medicine, Tribhuvan University; Dr. Meeta Singh, Department of Gynecology and Obstetrics, Nepalgunj Medical College; Dr. Ram Krishna Chandyo, Department of Community Medicine, Kathmandu Medical College; and Dr. Chakra Budhathoki, Johns Hopkins University School of Nursing. This study was conducted in collaboration with our implementing partner, Nepal Netra Jyoti Sangh, under the auspices of the Social Welfare Council of the Government of Nepal.

## Author Contributors

**Conceptualization:** Daniel J. Erchick, James M. Tielsch, Joanne Katz, Subarna K. Khatry, Parul Christian

**Data Curation**: Tsering P. Lama, Daniel J. Erchick

**Formal Analysis**: Tsering P. Lama

**Funding Acquisition:** Daniel J. Erchick

**Project Administration**: Subarna K. Khatry, Tsering P. Lama, Rita Shrestha, Steven C LeClerq

**Supervision**: Rita Shrestha, Tsering P. Lama

**Writing – original draft preparation**: Tsering P. Lama

**Writing – Review & Editing**: Tsering P. Lama, Rita Shrestha, Parul Christian, James M. Tielsch, Joanne Katz, Subarna K. Khatry, Steven C LeClerq, Daniel J. Erchick

## Data availability statement

Data described in the manuscript, code book, and analytic code will be made available upon reasonable request. Data can be requested from the research principal investigator

## Ethics

The study was approved by the Institutional Review Board (IRB) at Johns Hopkins Bloomberg School of Public Health, Baltimore, USA (IRB00009714), the Committee on Human Research IRB at The George Washington University, Washington, DC, USA (081739), and the Ethical Review Board of the Nepal Health Research Council, Kathmandu, Nepal (174/2018).

## Patient and public involvement

Patients and/or the public were not involved in the design, or conduct, or reporting, or dissemination plans of this research.

## Patient consent for publication

Not required.

## Trial registration number

NCT03668977.

## References

1. Lu C, Black MM, Richter LM. Risk of poor development in young children in low-income and middle-income countries: an estimation and analysis at the global, regional, and country level. Lancet Glob Health. 2016 Dec 1;4(12):e916–22. doi:10.1016/S2214-109X(16)30266-2 PubMed PMID: 27717632.

2. Schlotz W, Phillips DIW. Fetal origins of mental health: evidence and mechanisms. Brain Behav Immun. 2009 Oct;23(7):905–16. doi:10.1016/j.bbi.2009.02.001 PubMed PMID: 19217937.

3. Monk C, Georgieff MK, Osterholm EA. Maternal prenatal distress and poor nutrition – mutually influencing risk factors affecting infant neurocognitive development. J Child Psychol Psychiatry. 2013 Feb;54(2):115–30. doi:10.1111/jcpp.12000 PubMed PMID: 23039359; PubMed Central PMCID: PMC3547137.

4. Grantham-McGregor S, Cheung YB, Cueto S, Glewwe P, Richter L, Strupp B. Developmental potential in the first 5 years for children in developing countries. Lancet. 2007 Jan 6;369(9555):60–70. doi:10.1016/S0140-6736(07)60032-4/ATTACHMENT/2F933B60-1E68-4B3E-A529-97892C41953B/MMC2.PDF PubMed PMID: 17208643.

5. Kretchmer N, Beard JL, Carlson S. The role of nutrition in the development of normal cognition. Am J Clin Nutr. 1996 Jun;63(6):997S–1001S. doi:10.1093/ajcn/63.6.997 PubMed PMID: 8644701.

6. WHO recommendations on antenatal care for a positive pregnancy experience [Internet]. [cited 2026 May 21]. Available from: https://www.who.int/publications/i/item/9789241549912

7. Imdad A, Bhutta ZA. Effect of balanced protein energy supplementation during pregnancy on birth outcomes. BMC Public Health. 2011 Apr 13;11(SUPPL. 3):S17. doi:10.1186/1471-2458-11-S3-S17

8. Stevens B, Buettner P, Watt K, Clough A, Brimblecombe J, Judd J. The effect of balanced protein energy supplementation in undernourished pregnant women and child physical growth in low- and middle-income countries: A systematic review and meta-analysis. Matern Child Nutr. 2015 Oct 1;11(4):415–32. doi:10.1111/mcn.12183

9. Ota E, Hori H, Mori R, Tobe-Gai R, Farrar D. Antenatal dietary education and supplementation to increase energy and protein intake. Cochrane Database Syst Rev. 2015 Jun 2;2015(6):CD000032. doi:10.1002/14651858.CD000032.pub3

10. Members of an Expert Consultation on Nutritious Food Supplements for Pregnant and Lactating. Framework and specifications for the nutritional composition of a food supplement for pregnant and lactating women (PLW) in undernourished and low income settings. [Internet]. Gates OPen Res; 2019 [cited 2026 Mar 11]. Available from: https://gatesopenresearch.org/documents/3-1498doi:10.21955/gatesopenres.1116379.1

11. Ciulei M, Zhou S, Gallagher K, Taneja S, Bhandari N, Kolsteren P, et al. The effect of balanced energy-protein supplementation provided to lactating women on maternal and infant outcomes: study protocol for a prospectively planned individual patient data (IPD) meta-analysis [Internet]. F1000Research; 2024 [cited 2025 Jun 24]. Available from: https://f1000research.com/articles/13-598 doi:10.12688/f1000research.145501.1

12. Gernand AD, Gallagher K, Bhandari N, Kolsteren P, Lee AC, Shafiq Y, et al. Harmonization of maternal balanced energy-protein supplementation studies for individual participant data (IPD) meta-analyses – finding and creating similarities in variables and data collection. BMC Pregnancy Childbirth. 2023 Feb 11;23(1):107. doi:10.1186/s12884-023-05366-2

13. Muhammad A, Shafiq Y, Nisar MI, Baloch B, Pasha A, Yazdani NS, et al. Effect of maternal postnatal balanced energy protein supplementation and infant azithromycin on infant growth outcomes: an open-label randomized controlled trial. Am J Clin Nutr. 2024 Sep;120(3):550–9. doi:10.1016/j.ajcnut.2024.06.008 PubMed PMID: 38925354; PubMed Central PMCID: PMC11393397.

14. Argaw A, Kok B de, Toe LC, Hanley-Cook G, Dailey-Chwalibóg T, Ouédraogo M, et al. Fortified balanced energy–protein supplementation during pregnancy and lactation and infant growth in rural Burkina Faso: A 2 × 2 factorial individually randomized controlled trial. PLOS Med. 2023 Feb 6;20(2):e1004186. doi:10.1371/journal.pmed.1004186

15. Taneja S, Upadhyay RP, Chowdhury R, Kurpad AV, Bhardwaj H, Kumar T, et al. Impact of nutritional interventions among lactating mothers on the growth of their infants in the first 6 months of life: a randomized controlled trial in Delhi, India. Am J Clin Nutr. 2021 Apr 6;113(4):884–94. doi:10.1093/ajcn/nqaa383 PubMed PMID: 33564825; PubMed Central PMCID: PMC8023824.

16. Krebs NF, Lozoff B, Georgieff MK. Neurodevelopment: The Impact of Nutrition and Inflammation During Infancy in Low-Resource Settings. Pediatrics. 2017 Apr 1;139(Suppl 1):S50–8. doi:10.1542/PEDS.2016-2828G PubMed PMID: 28562248.

17. World Health Organization. The optimal duration of exclusive breastfeeding: Report of an expert consultation [Internet]. World Health Organization; 2001 Jan [cited 2025 Mar 25]. Available from: https://www.who.int/publications/i/item/WHO-NHD-01.09

18. Cusick SE, Georgieff MK. The Role of Nutrition in Brain Development: The Golden Opportunity of the “First 1000 Days.” J Pediatr. 2016 Aug 1;175:16. doi:10.1016/J.JPEDS.2016.05.013 PubMed PMID: 27266965.

19. Lönnerdal B. Effects of maternal dietary intake on human milk composition. J Nutr. 1986 Apr;116(4):499–513. doi:10.1093/jn/116.4.499 PubMed PMID: 3514820.

20. Larson LM, Yousafzai AK. A meta-analysis of nutrition interventions on mental development of children under-two in low- and middle-income countries. Matern Child Nutr. 2017 Jan;13(1):e12229. doi:10.1111/mcn.12229 PubMed PMID: 26607403; PubMed Central PMCID: PMC6866072.

21. Erchick DJ, Lama TP, Khatry SK, Katz J, Mullany LC, Zavala E, et al. Supplementation with fortified balanced energy–protein during pregnancy and lactation and its effects on birth outcomes and infant growth in southern Nepal: protocol of a 2×2 factorial randomised trial. BMJ Paediatr Open. 2023 Nov 3;7(1):2229. doi:10.1136/BMJPO-2023-002229 PubMed PMID: 10.1136/bmjpo-2023-002229.

22. Ministry of Health and Population (Nepal), New ERA, ICF. Nepal Demographic and Health Survey 2022. Kathmandu, Nepal: Ministry of Health and Population, Nepal; 2023.

23. Lama TP, Moore K, Isanaka S, Jones L, Bedford J, de Pee S, et al. Compliance with and acceptability of two fortified balanced energy protein supplements among pregnant women in rural Nepal. Matern Child Nutr. 2022 Apr 1;18(2):e13306. doi:10.1111/MCN.13306 PubMed PMID: 34908227.

24. Lama TP, Khatry SK, Isanaka S, Moore K, Jones L, Bedford J, et al. Acceptability of 11 fortified balanced energy-protein supplements for pregnant women in Nepal. Matern Child Nutr. 2022 Jul 1;18(3):e13336. doi:10.1111/MCN.13336 PubMed PMID: 35263004.

25. Bayley N, Aylward G. Bayley 4 Scales of Infant and Toddler Development, Fourth Edition; Technical Manual. Pearson; 2019.

26. Ranjitkar S, Kvestad I, Strand TA, Ulak M, Shrestha M, Chandyo RK, et al. Acceptability and Reliability of the Bayley Scales of Infant and Toddler Development-III Among Children in Bhaktapur, Nepal. Front Psychol. 2018 Jul 24;9(JUL). doi:10.3389/FPSYG.2018.01265 PubMed PMID: 30087639.

27. Murray-Kolb LE, Rasmussen ZA, Scharf RJ, Rasheed MA, Svensen E, Seidman JC, et al. The MAL-ED Cohort Study: Methods and Lessons Learned When Assessing Early Child Development and Caregiving Mediators in Infants and Young Children in 8 Low- and Middle-Income Countries. Clin Infect Dis Off Publ Infect Dis Soc Am. 2014 Nov 1;59(Suppl 4):S261. doi:10.1093/CID/CIU437 PubMed PMID: 25305296.

28. Shrestha PS, Shrestha SK, Bodhidatta L, Strand T, Shrestha B, Shrestha R, et al. Bhaktapur, Nepal: the MAL-ED birth cohort study in Nepal. Clin Infect Dis Off Publ Infect Dis Soc Am. 2014 Nov 1;59 Suppl 4:S300–303. doi:10.1093/cid/ciu459 PubMed PMID: 25305301.

29. Q-global Web-based Administration, Scoring, and Reporting [Internet]. [cited 2025 Apr 4]. Available from: https://www.pearsonassessments.com/en-us/Store/Professional-Assessments/Q-global-Web-based-Administration%2C-Scoring%2C-and-Reporting/p/100000680

30. Wijnhoven TM, de Onis M, Onyango AW, Wang T, Bjoerneboe GEA, Bhandari N, et al. Assessment of gross motor development in the WHO Multicentre Growth Reference Study. Food Nutr Bull. 2004 Mar;25(1 Suppl):S37–45. doi:10.1177/15648265040251S105 PubMed PMID: 15069918.

31. StataCorp. Stata statistical software: Release 15, College Station, TX. TX; 2017.

32. Upadhyay RP, Taneja S, Chowdhury R, Dhabhai N, Sapra S, Mazumder S, et al. Child Neurodevelopment After Multidomain Interventions From Preconception Through Early Childhood: The WINGS Randomized Clinical Trial. JAMA. 2024 Jan 2;331(1):28. doi:10.1001/JAMA.2023.23727 PubMed PMID: 38165408.

33. Sudfeld CR, Bliznashka L, Salifou A, Guindo O, Soumana I, Adehossi I, et al. Evaluation of multiple micronutrient supplementation and medium-quantity lipid-based nutrient supplementation in pregnancy on child development in rural Niger: A secondary analysis of a cluster randomized controlled trial. PLOS Med. 2022 May 2;19(5):e1003984. doi:10.1371/journal.pmed.1003984

34. Tofail F, Persson LÅ, Arifeen SE, Hamadani JD, Mehrin F, Ridout D, et al. Effects of prenatal food and micronutrient supplementation on infant development: a randomized trial from the Maternal and Infant Nutrition Interventions, Matlab (MINIMat) study. Am J Clin Nutr. 2008 Mar 1;87(3):704–11. doi:10.1093/AJCN/87.3.704 PubMed PMID: 18326610.

35. Christian P, Murray-Kolb LE, Khatry SK, Katz J, Schaefer BA, Cole PM, et al. Prenatal Micronutrient Supplementation and Intellectual and Motor Function in Early School-aged Children in Nepal. JAMA. 2010 Dec 22;304(24):2716–23. doi:10.1001/jama.2010.1861

36. Ouyang J, Cai W, Wu P, Tong J, Gao G, Yan S, et al. Association between Dietary Patterns during Pregnancy and Children’s Neurodevelopment: A Birth Cohort Study. Nutrients. 2024 May 19;16(10):1530. doi:10.3390/nu16101530 PubMed PMID: 38794768; PubMed Central PMCID: PMC11123670.

37. Zhang Z, Tran NT, Nguyen TS, Nguyen LT, Berde Y, Tey SL, et al. Impact of maternal nutritional supplementation in conjunction with a breastfeeding support program during the last trimester to 12 weeks postpartum on breastfeeding practices and child development at 30 months old. PLoS ONE. 2018 Jul 16;13(7):e0200519. doi:10.1371/journal.pone.0200519 PubMed PMID: 30011318; PubMed Central PMCID: PMC6047798.

38. Leung BMY, Wiens KP, Kaplan BJ. Does prenatal micronutrient supplementation improve children’s mental development? A systematic review. BMC Pregnancy Childbirth. 2011 Feb 3;11:12. doi:10.1186/1471-2393-11-12 PubMed PMID: 21291560; PubMed Central PMCID: PMC3039633.

39. Ip P, Ho FKW, Rao N, Sun J, Young ME, Chow CB, et al. Impact of nutritional supplements on cognitive development of children in developing countries: A meta-analysis. Sci Rep. 2017 Sep 6;7(1):10611. doi:10.1038/s41598-017-11023-4

40. Christian P, Kim J, Mehra S, Shaikh S, Ali H, Shamim AA, et al. Effects of prenatal multiple micronutrient supplementation on growth and cognition through 2 y of age in rural Bangladesh: the JiVitA-3 Trial12. Am J Clin Nutr. 2016 Oct 1;104(4):1175–82. doi:10.3945/ajcn.116.135178

41. Brito NH, Fifer WP, Amso D, Barr R, Bell MA, Calkins S, et al. Beyond the Bayley: Neurocognitive Assessments of Development During Infancy and Toddlerhood. Dev Neuropsychol. 2019;44(2):220–47. doi:10.1080/87565641.2018.1564310 PubMed PMID: 30616391; PubMed Central PMCID: PMC6399032.

42. Kvestad I, Hysing M, Ranjitkar S, Shrestha M, Ulak M, Chandyo RK, et al. The stability of the Bayley scales in early childhood and its relationship with future intellectual abilities in a low to middle income country. Early Hum Dev. 2022 Jul;170:105610. doi:10.1016/j.earlhumdev.2022.105610 PubMed PMID: 35728398.

43. Schonhaut L, Pérez M, Armijo I, Maturana A. Comparison between Ages & Stages Questionnaire and Bayley Scales, to predict cognitive delay in school age. Early Hum Dev. 2020 Feb 1;141:104933. doi:10.1016/j.earlhumdev.2019.104933

44. Thomas S, Thomas T, Bosch RJ, Ramthal A, Bellinger DC, Kurpad AV, et al. Effect of Maternal Vitamin B12 Supplementation on Cognitive Outcomes in South Indian Children: A Randomized Controlled Clinical Trial. Matern Child Health J. 2019 Feb 1;23(2):155–63. doi:10.1007/s10995-018-2605-z

45. Black MM, Fernandez-Rao S, Nair KM, Balakrishna N, Tilton N, Radhakrishna KV, et al. A Randomized Multiple Micronutrient Powder Point-of-Use Fortification Trial Implemented in Indian Preschools Increases Expressive Language and Reduces Anemia and Iron Deficiency. J Nutr. 2021 Jul 1;151(7):2029–42. doi:10.1093/jn/nxab066

46. Brazelton TB, Nugent JK. Wiley.com [Internet]. 2011 [cited 2025 Nov 13]. Neonatal Behavioral Assessment Scale, 4th Edition | Wiley. Available from: https://www.wiley.com/en-us/Neonatal+Behavioral+Assessment+Scale%2C+4th+Edition-p-9781907655036

47. Pence Turnbull KL, Justice LM. Language Development From Theory to Practice [Internet]. 3rd edition. Upper Saddle River, NJ: Pearson Education; 2017 [cited 2025 Nov 13]. Available from: https://www.pearson.com/en-us/subject-catalog/p/language-development-from-theory-to-practice/P200000001676/9780134170671?srsltid=AfmBOoo0M69uei9q5t5ycRYvU1nupVQGws5U_7mJOgp2vRPY8rEcvyB2

48. ASHA. American Speech-Language-Hearing Association [Internet]. American Speech-Language-Hearing Association; 2008 [cited 2025 Nov 13]. Roles and Responsibilities of Speech-Language Pathologists With Respect to Reading and Writing in Children and Adolescents. Available from: www.asha.org/policy/ps2001-00104

49. Prado EL, Dewey KG. Nutrition and brain development in early life. Nutr Rev. 2014 Apr 1;72(4):267–84. doi:10.1111/nure.12102

50. Georgieff MK. Nutrition and the developing brain: nutrient priorities and measurement2. Am J Clin Nutr. 2007 Feb 1;Maternal Nutrition and Optimal Infant Feeding Practices 85(2):614S-620S. doi:10.1093/ajcn/85.2.614S

51. Allen LH. Multiple micronutrients in pregnancy and lactation: an overview2. Am J Clin Nutr. 2005 May 1;81(5):1206S–1212S. doi:10.1093/ajcn/81.5.1206

52. Innis SM. Impact of maternal diet on human milk composition and neurological development of infants. Am J Clin Nutr. 2014 Mar;99(3):734S–41S. doi:10.3945/ajcn.113.072595 PubMed PMID: 24500153.

53. Dekaban AS. Changes in brain weights during the span of human life: relation of brain weights to body heights and body weights. Ann Neurol. 1978 Oct;4(4):345–56. doi:10.1002/ana.410040410 PubMed PMID: 727739.

54. Couperus JW, Nelson CA. Early Brain Development and Plasticity. In: Blackwell Handbook of Early Childhood Development [Internet]. John Wiley & Sons, Ltd; 2006 [cited 2025 Nov 11]. p. 85–105. Available from: https://onlinelibrary.wiley.com/doi/abs/10.1002/9780470757703.ch5 doi:10.1002/9780470757703.ch5

55. Grossman AW, Churchill JD, McKinney BC, Kodish IM, Otte SL, Greenough WT. Experience effects on brain development: possible contributions to psychopathology. J Child Psychol Psychiatry. 2003 Jan;44(1):33–63. doi:10.1111/1469-7610.t01-1-00102 PubMed PMID: 12553412.

56. Thompson RA, Nelson CA. Developmental science and the media. Early brain development. Am Psychol. 2001 Jan;56(1):5–15. doi:10.1037/0003-066x.56.1.5 PubMed PMID: 11242988.

57. Britto PR, Lye SJ, Proulx K, Yousafzai AK, Matthews SG, Vaivada T, et al. Nurturing care: promoting early childhood development. The Lancet. 2017 Jan 7;389(10064):91–102. doi:10.1016/S0140-6736(16)31390-3

58. Aboud FE, Yousafzai AK. Global Health and Development in Early Childhood. Annu Rev Psychol. 2015 Jan 3;66(Volume 66, 2015):433–57. doi:10.1146/annurev-psych-010814-015128

59. Prado EL, Larson LM, Cox K, Bettencourt K, Kubes JN, Shankar AH. Do effects of early life interventions on linear growth correspond to effects on neurobehavioural development? A systematic review and meta-analysis. Lancet Glob Health. 2019 Oct 1;7(10):e1398–413. doi:10.1016/S2214-109X(19)30361-4

60. Zhu Z, Chang S, Cheng Y, Qi Q, Li S, Elhoumed M, et al. Early life cognitive development trajectories and intelligence quotient in middle childhood and early adolescence in rural western China. Sci Rep. 2019 Dec 4;9(1):18315. doi:10.1038/s41598-019-54755-1 PubMed PMID: 31797987; PubMed Central PMCID: PMC6892923.

61. Colombo J. Assessing Neurocognitive Development in Studies of Nutrition. In: Nestle Nutrition Institute workshop series. Basel, Switzerland: S. Karger AG; 2018. p. 143–54. (Recent Research in Nutrition and Growth). doi:10.1159/000486499

62. Jiang T, Christian P, Khatry SK, Wu L, West KP. Micronutrient Deficiencies in Early Pregnancy Are Common, Concurrent, and Vary by Season among Rural Nepali Pregnant Women1. J Nutr. 2005 May 1;135(5):1106–12. doi:10.1093/jn/135.5.1106

